# Epidemiological Profile of Crimean-Congo Hemorrhagic Fever, Iraq, 2018

**DOI:** 10.1101/2023.11.22.23298722

**Authors:** Ali Hazim Mustafa, Faris Lami, Hanan Abdulghafoor Khaleel

## Abstract

**Background:** Crimean-Congo hemorrhagic fever (CCHF) is a potentially fatal tick-borne disease that is widely distributed in Africa and Eurasia countries. It is caused by the CCHF virus of the *Nairovirus* genus of the *Bunyaviridae* family.

This study aims to describe the 2018 CCHF epidemic wave in Iraq and epidemiological pattern to assist implantation of preventive and control measures and adherence of physicians to the standard case definition.

**Methods:** This descriptive study reviewed all records of suspected and confirmed CCHF cases. Three types of data sources were used: the case investigation forms of all suspected cases, case sheets of all confirmed cases, and the laboratory results from the central public health laboratory.

**Results:** The total number of suspected cases was 143. Most of the cases were males (59.4%), 15-45 years old (62.2%), and live in urban areas (58.7%). About three quarters of the cases (68.5%) did not fit the standard case definition adopted by Iraq Center of Disease Control. Most of the suspected cases were reported in Diwaniya province (20.3%). Nearly half of them (64, 44.7%) occurred in June.

Only 7.0% (n=10) of suspected cases were positive when tested by Reverse Transcriptase Polymerase Chain Reaction (RT-PCR). One third of confirmed cases (3, 30.0%) occurred in Diwaniya province. During the 2018 epidemic wave, there were 10 confirmed cases with 8 deaths and 2 improved cases.

**Conclusion:** Despite the fact that CCHF is uncommon in Iraq, sporadic cases or outbreaks could occur.

**Recommendations:** Given the known method of transmission, banning of random livestock slaughtering and the practice of raising livestock inside residential areas are expected to have a major role in CCHF infection control.

## 1. Introduction

Crimean-Congo hemorrhagic fever (CCHF) virus is a member of the *Nairovirus* genus family *Bunyaviridae* [1]. CCHF virus is a zoonotic agent causing severe life-threatening diseases with a case fatality rate of 10-50% [2]. The distribution of the disease coincides with geographical occurrence of its tick vector *Haylomma Marginatum* [3]., Human infection occurs through tick bites [4], crushed infected ticks, contact with patients infected with CCHF during the acute phase of infection, or contact with blood or tissues from viraemic livestock [5]. CCHF is also an occupational disease for butchers, slaughterhouse workers, livestock workers, animal husbandry workers, veterinarians, and health care workers [6]., Additionally, living in rural areas is a risk factor for tick exposure and thus, for acquiring infection [7].

The incubation period of the virus is 1-7 days [8]. The first sign of CCHF is a sudden onset of very high fever with nonspecific symptoms [9]. Rapid diagnosis is achieved by (RT-PCR) test [10]. There is no proven vaccine for human use or animal use [11], and treatment is mainly supportive [12]. CCHF was recognized for the first time in Iraq in 1979 [13], however, the literatures on CCHF in Iraq is scarce Accordingly.

The aim to identify epidemiological pattern of the disease in Iraq by reviewing all medical case reports of suspected and confirmed cases of the 2018 epidemic wave to inform on the proper implementation of the preventive and control measures and adherence of physicians to the standard case definition to curb this viral infection.

## 2. Methods

This descriptive study is based on a retrospective records review of hospitals and directorates of health in Iraq provinces as well as the Medical City Directorate of Health in Baghdad. Accordingly, a suspected case of CCHF was defined as acute onset of fever, hemorrhagic symptoms, and a history of animal contact; and a confirmed case was defined as a suspected case with serologic confirmation using enzyme-linked immunosorbent assay (ELISA) and/or PCR tests.

Three types of data sources were used, the first included case investigation forms for all suspected cases that were received at the zoonotic disease section of the Iraq Center of Disease Control (CDC). The second included case sheets of all confirmed cases who were admitted to the hospitals in all Iraqi governorates. These case sheets were retrieved from the hospitals after granting approvals from all concerned bodies at the central and provincial levels. The third data source included laboratory results received from the Central Public Health Laboratory (CPHL) for ELISA and/or RT-PCR tests. All blood samples from suspected cases were tested at the Central Public Health Laboratory/ Virology Department /CCHF laboratory, by using the advanced RT- PCR Test.

A check-list to do record review was developed to collect information from the suspected and confirmed cases. The variables included in the case investigation forms were selected based on case definition adopted by Iraq CDC.

The variables included in the case investigation forms were selected based on case definition adopted by Iraq CDC. The variables collected include the reporting directorate of health, time of reporting, primary diagnosis, demographic characteristics of patients, main signs and symptoms, date of symptoms onset, investigation results, history of exposure, and risk factors including (- residency in rural area, tick exposure, contact with animals, direct contact with uncooked meat, contact with similar case, and occupation).

- Official approval was granted from the Ministry of Health (MOH)/Directorate of Public Health
- All data were kept confidential and used exclusively for the sake of the research.

### Statistical Analysis

Frequency and percentage (n, %) were used to describe the distribution of continuous variables. A detailed description of each confirmed case was presented, which include all relevant epidemiological, clinical, laboratory, and management details. Epi-info 7 program was used for data entry and analysis.

## 3. Results

### a. Suspected CCHF cases

In 2018, a total of 143 suspected CCHF cases were identified from January 31st 2018, as the date of symptoms onset of the first case, till November 18th, 2018, which is the date of symptoms onset of the last case. Most cases were males (59.4%), between the ages of (15 and 45) (62.2%), and living in urban areas (58.7%). Among the high-risk group, only 11 cases (7.7%) were animal owner and (3.5%) were butchers, Table 1.

**Table 1:** Characteristics of suspected and confirmed CCHF cases.

| **Characteristics** | **Suspected cases (N, %)** | **Confirmed cases (N, %)** |
| --- | --- | --- |
| **Total** | **143** | **10** |
| **Age** |  |  |
| < 5 yr. | 9 (6.3) | 1 (10) |
| 5– 14 yr. | 21(14.7) | 1 (10) |
| 15 – 45 yr. | 89 (62.2) | 5 (50) |
| >45 yr. | 24 (16.8) | 3 (30) |
| **Sex** |  |  |
| Male | 85 (59.4) | 9 (90) |
| Female | 58 (40.6) | 1 (10) |
| **Residency** |  |  |
| Urban | 84 (58.7) | 1 (10) |
| Rural | 56 (39.2) | 9 (90) |
| Displaced camps | 3 (2.1) |  |
| **Occupation** |  |  |
| Other | 61 (42.7) | 1 (10) |
| House wife | 30 (21.0) | 0 |
| Child | 21 (14.7) | 1 (10) |
| Animal owner | 11 (7.7) | 4 (40) |
| Farmer | 6 (4.2) | 1 (10) |
| Health worker | 6 (4.2) | 0 |
| Butcher | 5 (3.5) | 3 (30) |
| Military | 3 (2.1) | 0 |
| **Fitness to case definition** | 45 (31.5) | 10(100) |
| **Main signs and symptom** |  |  |
| Fever | 91 (63.6) | 10 (100) |
| Bleeding from Body openings | 45 (21.11) | 2 (20) |
| Ecchymosis | 25 (11.7) | 6 (60) |
| Gum bleeding | 21 (9.9) | 9 (90) |
| Injection sites bleeding | 16 (7.5) | 1 (10) |
| Congested conjunctiva | 15 (7) | 0 |

According to the case investigation forms of all suspected cases that were finally received at the zoonotic disease section/ CDC/ Iraq, only (31.5%) fit to the standard case definition, and (68.5%) of suspected cases did not fit the case definition, but relayed only on one symptom, which was either fever, or presence of hemorrhagic symptoms, with or without a history of animal contact. In addition, there was a lack of accuracy in filling several fields in the check-list forms including (risk factors, clinical signs and symptoms, educational qualification, and socioeconomic status).

Cases were reported in all Iraq provinces except Sulaymaniya province, The provinces reporting the most CCHF suspected cases were Diwaniya (20.3%), Babylon (14.7%), and Basra (11.9%). During this outbreak, 10 cases out of 143 cases were RT-PCR positive when tested in CPHL in Baghdad, Table 2.

**Table 2:** The distribution of suspected and confirmed cases by the reported DOH.

| Reported DOH | Suspected cases (N, %) | Confirmed cases (N, %) |
| --- | --- | --- |
| **Total** | **143** | **10** |
| DIWANIYA | 29 (20.3) | 3 (30.0) |
| BABYLON | 21 (14.7) | 0 |
| BASRA | 17 (11.9) | 1 (10) |
| BAGHDAD-RESAFA | 12 (8.4) | 1 (10) |
| ERBIL | 11 (7.7) | 2 (20) |
| BAGHDAD-KARKH | 8 (5.6) | 0 |
| DIYALA | 8 (5.6) | 0 |
| THI-QAR | 7 (4.9) | 0 |
| NINEWA | 7 (4.9) | 2 (20) |
| NAJAF | 6 (4.2) | 1 (10) |
| DAHUK | 4 (2.8) | 0 |
| WASSIT | 3 (2.1) | 0 |
| MISSAN | 2 (1.4) | 0 |
| MUTHANNA | 2 (1.4) | 0 |
| MEDICAL CITY | 2 (1.4) | 0 |
| SALAH AL-DIN | 2 (1.4) | 0 |
| KARBALA | 1 (0.7) | 0 |
| KIRKUK | 1 (0.7) | 0 |

**Figure 1.**
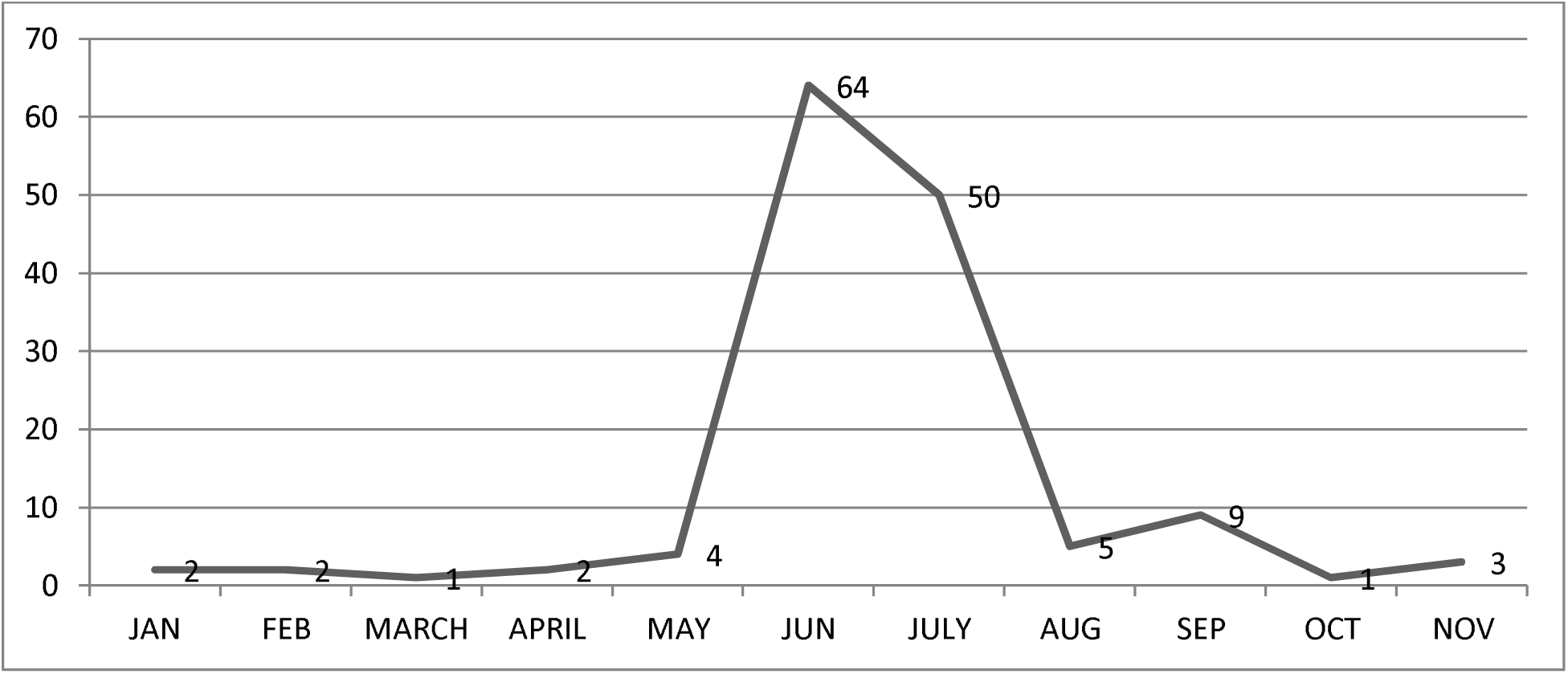
The number of cases increased in the months of summer especially in June and July.

Sporadic cases were reported in winter and spring during the period from January till May,

The information provided on the history of exposure revealed that 11.9% of the suspected cases had positive history of contact with a confirmed case, about 30.8% have a history of animal contact, tick exposure was reported in 19.6% of the cases, and direct contact with uncooked meat was reported in 14.7% of the cases, Table 3.

**Table 3:** Distribution of suspected and confirmed CCHF cases according to risk factors and history of exposure.

| Risk factor | Suspected cases N (%) | Confirmed cases N (%) |
| --- | --- | --- |
| **Total** | **143** | **10** |
| Contact with similar case | 17 (11.9) | 0 |
| Contact with animals | 44 (30.8) | 5 (50) |
| Uncooked meat contact | 21 (14.7) | 5 (50) |
| Tick exposure | 28 (19.6) | 3 (30) |
| Slaughtering at home | 12 (8.4) | 0 |
| Residency in rural areas | 56 (38.5) | 9 (90) |
| Butcher | 5 (3.5) | 3 (30) |

### b. The Confirmed CCHF cases

Table 4 provides an in-depth description of the 10 confirmed cases of CCHF. Nine cases were males and only one case was a female. By occupation, four cases occurred among animal owners and three cases among butchers. Confirmed cases of CCHF were reported in different provinces in Iraq. By residency, four cases occurred in Diwaniya, two in Erbil, two in Ninewa 2, and one each in Thi-Qar and Diyala. Accordingly, most confirmed cases were seen in Diwaniya governorate (30%). The vast majority were residents of rural areas, 50% had contact with animals and 50% had contact with uncooked meat.

**Table 4:** In-depth description of the 10 confirmed CCHF cases reported in Iraq in 2018.

| Case ID | Age (years ) | Sex | Residence | Occupation | Risk Factors | Major signs and symptoms | Investigations | Outcome |
| --- | --- | --- | --- | --- | --- | --- | --- | --- |
| 1 | 60-69 | Male | rural | Farmer | Animal owner | Fever, Gum and GIT bleeding, DLOC. | PLT↓.<br>↑PT, PTT, INR.<br>↑liver enzymes. | Death |
| 2 | 20-29 | Male | rural | Student | Animal owner | Fever, Gum and GIT Bleeding, Epistaxis, Ecchymosis. | PLT↓.<br>↑PT, PTT, INR.<br>↑liver enzymes<br>↑ WBC | Death |
| 3 | 30-39 | Male | rural | Worker | Animal owner | Fever, Gum bleeding, ecchymosis, agitation, liver failure. | PLT↓.<br>↑PT, PTT, INR.<br>↑TSB, Blood Urea.<br>↑liver enzymes. | Death |
| 4 | 30-39 | Male | rural | Butcher | Contact with animal blood and tissues. | Fever, bleeding gums, ecchymosis, lymphadenopathy. | Positive ELISA and RT- PCR<br>↑TSB and liver Enzymes<br>↑blood urea | Discharged Well |
| 5 | 20-29 | Male | urban | Butcher | Contact with animal blood and tissues. | Fever, bleeding from body openings. | Positive RT-PCR test.<br>↑liver Enzymes<br>↑ALP<br>↑WBC. | Death |
| 6 | 10-19 | Male | rural | Animals owner | Contact with animals. | Fever, hematemesis, malena, bleeding from body openings, lymphadenopathy, confusion. | ↓PLT.<br>↑(PT, PTT, INR).<br>↑TSB and Liver Enzymes<br>↑LDH,<br>↓Total Protein | Discharged well |
| 7 | 0-9 | Male | rural | Child | Contact with animals and ticks. | Fever, Gum bleeding, Bleeding from injection sites, hematemesis, malena, epistaxis. | Positive ELISA and RT-PCR.<br>↓PLT.<br>↑PT, PTT, INR.<br>↑liver enzymes.<br>↓hemoglobin. | Death |
| 8 | 60-69 | Male | rural | Worker | Contact with animals. | Fever, bleeding gums, abdominal pain, malena. | Positive RT-PCR test.<br>↓PLT.<br>↑PT, PTT, INR.<br>↑S. AST (GOT). | Death |
| 9 | 40-49 | Male | rural | Butcher | Contact with animals' tissues and blood. | Fever, bleeding gums. ecchymosis, abdominal pain. | Positive RT-PCR test.<br>↓PLT.<br>↑PT, PTT, INR.<br>↑ALP, ↓Hemoglobin.<br>↑TSB and liver Enzymes.<br>↓WBC | Death |
| 10 | 50-59 | Female | rural | Animal Owner. | Contact with animals' tissues and blood. | Fever, bleeding gums. | No information available about biochemical tests and the treatment outlines. This case admitted to private hospital in ERBIL province. | Death |

**Abbreviations:**

Table5. The most commonly reported complication was altered level of consciousness, Figure 2, and outcome data revealed improvement of two of the 10 confirmed cases while eight deaths occurred after admission to a hospital.

**Figure 3:**
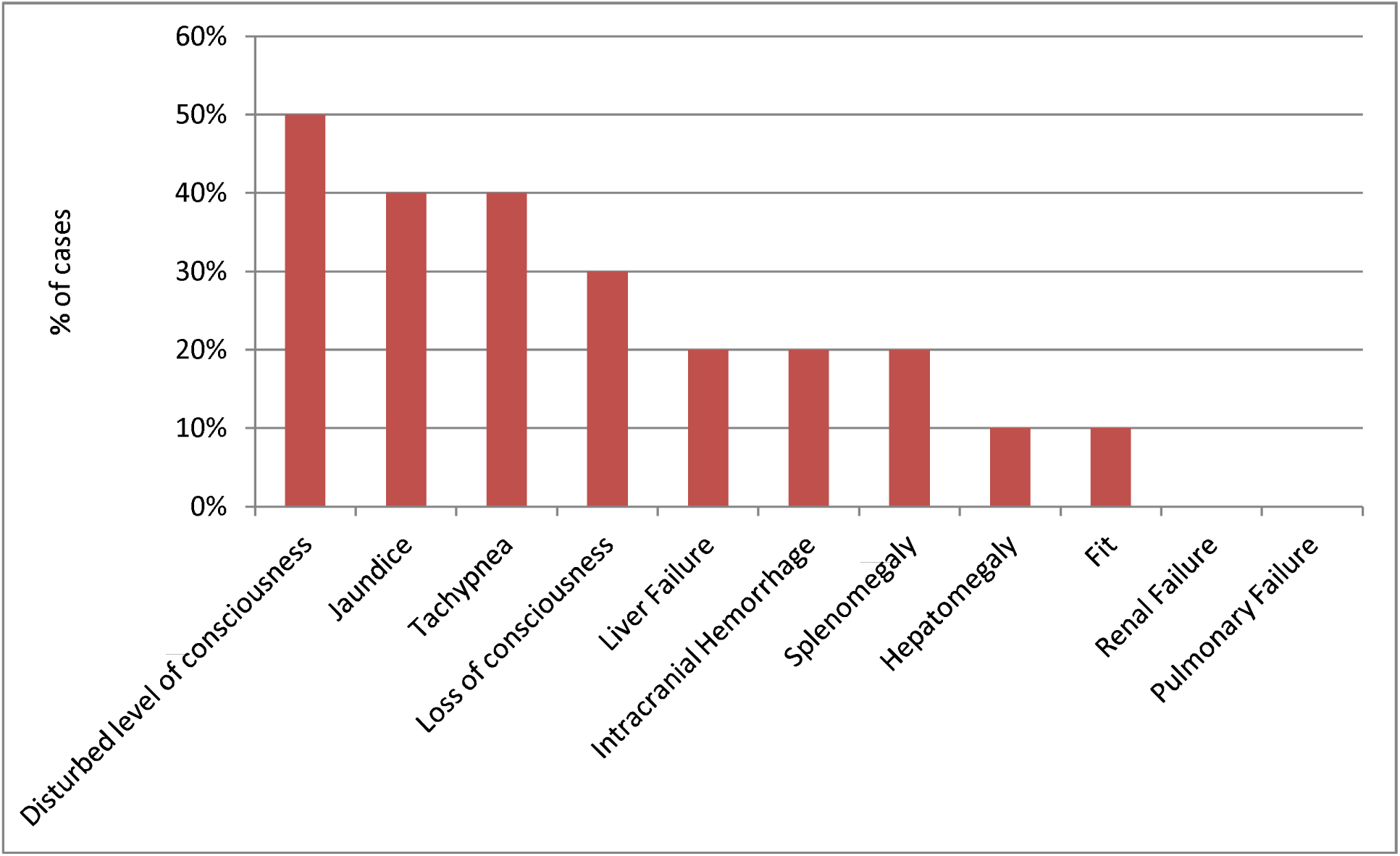
The main complications of confirmed CCHF cases.

**Table 5:** The treatment outlines of confirmed CCHF cases.

| Case ID | Ribavirin Treatment | Antibiotics | Number of transfusion units received. |  |  |  | Respiratory Support | Intravenous Fluid |
| --- | --- | --- | --- | --- | --- | --- | --- | --- |
|  |  |  | Blood | PLT. | FFP | CRYO |  |  |
| 1 | Not Used | Ceftriaxone .<br>Vancomycin .<br>Ampicillin . | 3 | 5 | 6 | 0 | Admission to ICU. | Normal saline 2000 cc. |
| 2 | Not Used | Claforan. | 4 | 10 | 10 | 10 | Admission to RCU. | Glucose water 2000cc.<br>Glucose saline 500 cc. |
| 3 | Not Used | Gentamycin.<br>Ceftriaxone.<br>Ciprodar .<br>Meronium .<br>Vacomycin . | 3 | 10 | 10 | 10 | SPO2 monitoring. | Normal saline 1500cc.<br>Ringer lactate 1500cc. |
| 4 | Not Used | Claforan .<br>Azithromycin. | 3 | 2 | 2 | 0 | Not Used. | Glucose water2000cc.<br>Glucose saline 2000cc. |
| 5 | Not Used | Ceftriaxone. | 0 | 1 | 0 | 0 | Not Used | Glucose saline |
| 6 | Used | Ceftriaxone. | 4 | 0 | 4 | 0 | Oxygen PRN. | Ringer lactate 1000cc.<br>Glucose saline 1000cc. |
| 7 | Not Used | Amoxil.<br>Claforan. | 1 | 1 | 1 | 0 | Not Used | 1/5 Glucose saline<br>750 cc/ 24 hours. |
| 8 | Not Used | Meronium. | 4 | 20 | 13 | 0 | Oxygen<br>SPO2 monitoring | Ringer lactate 1x6.<br>Normal saline 2000cc. |
| 9 | Used | Meronium .<br>Ciprodar. | 5 | 20 | 10 | 0 | Oxygen.<br>SPO2 monitoring. | Glucose saline1500cc<br>Normal saline1000cc |
| 10 | No information available about the treatment outlines of this case. She was admitted to private hospital in ERBIL province. Haemodialysis was not used in any of the cases. |  |  |  |  |  |  |  |
| **Abbreviations:**<br>**PLT:** platelets. **FFP:** Fresh Frozen Plasma. **Cryo:** cryoprecipitate. **PRN:** Pro re nata. |  |  |  |  |  |  |  |  |

## 4. Discussion

This study describes CCHF cases that were reported in Iraq in 2018. New suspected cases were in all Iraq provinces except Sulaimaniya governorates in Kurdistan region, and Diwaniya province saw the highest number of both suspected and confirmed cases. Most confirmed cases were males living in rural areas, and most were exposed to either uncooked meat or to animals. The main CCHFV risk factors reported in confirmed cases such as living in villages, occupational exposure, being an animal owner or working as a butcher, and positive history of tick bites were similar to those in other countries [14].

In 1996, tick control campaigns in Iraq results in a decrease in annual number of CCHF cases. Between 1998 and 2009 the annual number of cases ranged from zero to six, while in 2010 they increased to 11 due to lack of compliance with tick control activities. This reflects the actual need of regular laboratory investigation for the presence of infection in livestock and the frequent checking for the extent of livestock tick infestation with application of the suitable immediate preventive measures such as cattle and barn spraying with sheep and goat dipping [15].

No cases were reported in Sulaimaniya governorate since 1997 due to better eradication of ticks which are the reservoir and vector of the virus implemented by the veterinary authorities in the province This was proven in a serological and molecular study that was conducted in 2016, where blood samples collected from human and cattle, including butchers working at a slaughterhouse, tested negative on RT-PCR. Ticks were also collected for virus detection from three villages in the Sharazoor district of Sulaimaniya province. The tissue prepared from ticks also was negative for CCHFV [16].

Diwaniya DOH was the highest in the reporting of both suspected (20.3%) and confirmed cases (30.0%). This is may be related to working in agriculture and animal husbandry in rural areas [17] and lack of vector control [18] or the over reporting due to apprehension and concern after the first confirmed case death in the province. The reporting of suspected cases was much higher in urban areas (58.7%) than in rural areas (38.5%). This could be related to keeping livestock inside houses or residential areas by dwellers of urban areas or urban areas with rural culture. Also, the presence of special markets for animal trading in the outskirts of large cities might increase the chance of contact of resident with infected animals, Occasional contact with livestock could be effective in transmission of the virus [19]. Although tick exposure is one of the most important risk factors for CCHF, it cannot explain all cases and there are other important risk factors such as high-risk occupations. Even taking care of livestock for a short period at home can increase the chance of acquiring CCHF virus infection. The main route of transmission in Iraq is handling livestock in its viremic period [20].

The majority of confirmed cases occurred in summer and this is in accordance with activation of transmitting vector in temperate areas in late spring and continuous activation through summer to early autumn [21].

The standard case definition of suspected cases in Iraq is based on the WHO standard case definition, and this was distributed to medical staff through the annual plan of zoonotic diseases section [22]. However, still 69% of the total suspected cases did not fit the standard case definition and the medical staff relied mostly on one symptom, either fever or bleeding with or without a satisfactory history of animal contact.

Supportive treatment and management of hemodynamic status were essential in the treatment of confirmed CCHF cases and they were accordingly initiated on admission [23]. Similarly, the follow up of each patient was determined by individual clinical status and laboratory results, as is recommended in the literature [24].

Although the efficacy of ribavirin in the treatment of CCHF has not been proven conclusively, its use in the early stage of the disease is recommended [25]. Ribavirin, however, was only used in the treatment of two confirmed cases and no obvious reason could be discerned why it was not used in the other cases despite its availability. Corticosteroids treatment was used in combination with Ribavirin in six patients, to increase platelet count and reduced the need for blood products particularly in severe cases, and this is supported by evidence from the literature [26].

The case fatality rate was 80%, and half of these cases dwell in Diwaniya governorate. The cause of that may be related to the non-use of antiviral treatment or late presentation of cases [27]. Both sporadic and epidemic forms have been documented in regions of Africa, Asia, and Eastern Europe with case fatality rates ranging from 13 – 90% [28]. The number of infected and fatal cases reported to the Monitoring Emerging Diseases ProMED program between 1998 and 2013 showed that most of the cases were reported from Turkey, followed by Russia, Iran, Pakistan, and Afghanistan. The lowest fatalities were reported in Russia (4%) and Turkey (10%). Case fatality rates in other countries are as follows: Iran (12%), Pakistan (40%), Afghanistan (26%), Kazakhstan (39%), India (60%), Tajikistan (82%), United Arab Emirates (40%), Oman (100%) [29].

In response to the 2018 outbreak, preventive measures were taken by the Iraq MOH, including increasing health awareness among the population, medical care for suspected and confirmed cases admitted to hospitals, taking precautionary measures to prevent hospital acquired infections, follow up of contacts, and direct supervision of safe burial [30]. In addition to the coordination with Ministry of Agriculture – The General Company of Veterinary Affairs for the implementation of national awareness and livestock ticks control campaigns in animals by sheep and goat dipping and barn and cattle spraying with insecticides.

## 5. Conclusion

Although uncommon, the number of cases of CCHF increases in 2018, and they were detected all over Iraq. However, gaps remain in the proper use of standard case definitions and in certain cases of managements. CCHF poses a growing threat, and this is a call for action for policy makers to implement the needed preventive and control measures to curb the spread of this virus.

## 6. Recommendations

The current CCHF epidemiology in Iraq warrants continuing the health education and awareness campaigns targeted towards the public and medical society about the important preventive measures of the disease as well as of the signs and symptoms of the disease.

In addition, there is a need to enhance the level of coordination and cooperation with the veterinary company of the Ministry of Agriculture about the livestock tick control campaigns.

Other ministries and relevant authorities need to take the necessary measures and to prevent the phenomenon of random slaughtering of livestock outside the licensed government slaughterhouses as well as prevent raising livestock inside residential areas and inside houses.

All DsoH in Iraq have to strictly follow the WHO case definition to diagnose CCHF cases, set up a surveillance system all over the country to report any suspected case.

, conduct sero-prevalence studies among high-risk groups and hotspots, and use the Epi-info 7 form in the reporting of all suspected and confirmed cases of CCHF and pay attention to the extent of documentation within the fields of the form.

## Data Availability

All data produced in the present study are available upon reasonable request to the authors

